# Loss-of-function variants in *CUL3* cause a syndromic neurodevelopmental disorder

**DOI:** 10.1101/2023.06.13.23290941

**Authors:** Patrick R. Blackburn, Frédéric Ebstein, Tzung-Chien Hsieh, Marialetizia Motta, Francesca Clementina Radio, Johanna C. Herkert, Tuula Rinne, Isabelle Thiffault, Michele Rapp, Mariel Alders, Saskia Maas, Bénédicte Gerard, Thomas Smol, Catherine Vincent-Delorme, Benjamin Cogné, Bertrand Isidor, Marie Vincent, Ruxandra Bachmann-Gagescu, Anita Rauch, Pascal Joset, Giovanni Battista Ferrero, Andrea Ciolfi, Thomas Husson, Anne-Marie Guerrot, Carlos Bacino, Colleen Macmurdo, Stephanie S. Thompson, Jill A. Rosenfeld, Laurence Faivre, Frederic Tran Mau-Them, Wallid Deb, Virginie Vignard, Pankaj B. Agrawal, Jill A. Madden, Alice Goldenberg, François Lecoquierre, Michael Zech, Holger Prokisch, Ján Necpál, Robert Jech, Juliane Winkelmann, Monika Turčanová Koprušáková, Vassiliki Konstantopoulou, John R. Younce, Marwan Shinawi, Chloe Mighton, Charlotte Fung, Chantal Morel, Jordan Lerner- Ellis, Stephanie DiTroia, Magalie Barth, Dominique Bonneau, Ingrid Krapels, Sander Stegmann, Vyne van der Schoot, Theresa Brunet, Cornelia Bußmann, Cyril Mignot, Thomas Courtin, Claudia Ravelli, Boris Keren, Alban Ziegler, Linda Hasadsri, Pavel N. Pichurin, Eric W. Klee, Katheryn Grand, Pedro A. Sanchez-Lara, Elke Krüger, Stéphane Bézieau, Hannah Klinkhammer, Peter Michael Krawitz, Evan E. Eichler, Marco Tartaglia, Sébastien Küry, Tianyun Wang

## Abstract

**Purpose:** *De novo* variants in *CUL3* (Cullin-3 ubiquitin ligase) have been strongly associated with neurodevelopmental disorders (NDDs), but no large case series have been reported so far. Here we aimed to collect sporadic cases carrying rare variants in *CUL3,* describe the genotype-phenotype correlation, and investigate the underlying pathogenic mechanism.

**Methods:** Genetic data and detailed clinical records were collected via multi-center collaboration. Dysmorphic facial features were analyzed using GestaltMatcher. Variant effects on CUL3 protein stability were assessed using patient-derived T-cells.

**Results:** We assembled a cohort of 35 individuals with heterozygous *CUL3* variants presenting a syndromic NDD characterized by intellectual disability with or without autistic features. Of these, 33 have loss-of-function (LoF) and two have missense variants. *CUL3* LoF variants in patients may affect protein stability leading to perturbations in protein homeostasis, as evidenced by decreased ubiquitin-protein conjugates *in vitro*. Specifically, we show that cyclin E1 (CCNE1) and 4E-BP1 (EIF4EBP1), two prominent substrates of CUL3, fail to be targeted for proteasomal degradation in patient-derived cells.

**Conclusion:** Our study further refines the clinical and mutational spectrum of *CUL3*-associated NDDs, expands the spectrum of cullin RING E3 ligase-associated neuropsychiatric disorders, and suggests haploinsufficiency via LoF variants is the predominant pathogenic mechanism.

## Introduction

Cullin 3 (aka CUL3, or PHA2E) is an evolutionarily conserved component of the CUL3-RING E3 ubiquitin ligase (CRL) complex, which also includes a RING (really interesting new gene) -box protein, a Bric-a-brac/Tramtrack/Broad (BTB) protein, and other protein adaptors which together mediate the selective ubiquitination and subsequent proteasomal degradation of a variety of target proteins.^1,2^ The Cullin-RING Ligases (CRLs) include eight known highly conserved cullin protein members: CUL1, CUL2, CUL3, CUL4A, CUL4B, CUL5, CUL7 and CUL9.^1^ Germline variants in several of the genes encoding these proteins have been associated with syndromic neurodevelopmental disorders (NDDs), namely, X-linked syndromic intellectual disability (ID), Cabezas type (MRXSC, OMIM #300354) caused by hemizygous *CUL4B* variants,^3,4^ and 3-M syndrome (OMIM #273750) caused by biallelic *CUL7* variants.^5^

Heterozygous, predominantly *de novo*, gain-of-function variants affecting splicing of exon 9 in *CUL3* cause autosomal dominant pseudohypoaldosteronism type IIE (PHA2E, OMIM #614496), which presents with hyperkalemia and hyperchloremia with hyperchloremic metabolic acidosis leading to early-onset hypertension.^6^ Loss-of-function (LoF) variants in *CUL3* have also been reported in several large cohorts of patients with congenital heart defects, suggesting it may also have a broader role in early development.^7^ Trio-based exome sequencing studies in families with sporadic cases of autism spectrum disorder (ASD) have uncovered *de novo CUL3* variants in affected children (OMIM # 619239).^8–10^ Recently, *CUL3* was also found to be associated with an increased risk of ASD in large genome-wide association studies.^11,12^

Although *CUL3* is ubiquitously expressed in the brain, its role in the central nervous system is still poorly understood. Previous studies in mice have suggested that physical association of the potassium channel tetramerization domain containing-13 (Kctd13) protein, a substrate-specific adapter of the Cul3 ubiquitin-protein ligase complex, together helps mediate the ubiquitination of RhoA, leading to its degradation.^13^ Degradation of RhoA regulates the actin cytoskeleton as well as synaptic transmission. Modulation of this interaction within inner cortical plate layer 4 neurons may also influence corticogenesis and overall brain size in mice.^13^ ASD-associated nonsense variants in *CUL3* have been reported to disrupt the physical interaction between KCTD13 and CUL3, which may suggest a possible pathogenic mechanism for *CUL3* haploinsufficiency.^13^

Changes in neuronal migration and lamination defects have been previously associated with ASD in both human and mouse models, thus implicating *CUL3* haploinsufficiency in the development of ASD-related neurobehavioral abnormalities. More recently, Dong et al. found that *Cul3*^+/-^ mice showed social and behavioral abnormalities with increased anxiety levels.^14^ The hippocampal neurons were found to have increased spine density, neuronal excitability, synaptic transmission, and disrupted excitation-inhibition (E-I) balance in CA1 neurons. Pyramidal neurons also showed similar perturbations, demonstrating a role for Cul3 in synaptic dynamics including E-I balance.^14^ *Cul3* haploinsufficiency increased the level of Eif4g1, a protein involved in translational homeostasis, and upregulated Cap-dependent protein synthesis in the brains of mice, which may underlie the neurodevelopmental and neuropsychological deficits.^14^ In another study, Morandell et al.^15^ additionally showed that Cul3 is an important regulator of neuronal migration during a critical temporal window early in mouse development; *Cul3*^+/-^ mice developed cortical lamination abnormalities that manifested as motor and behavioral deficits.^15^ Loss of Cul3 led to upregulation of several proteins including plastin3 (Pls3), which was found to regulate actin organization and cell migration speed at a rate inversely proportional to its levels.^15^ This suggested that Cul3 may regulate cytoskeletal organization by controlling Pls3 levels in the brain and disrupt the protein homeostasis during a critical developmental window, which together results in the cortical changes observed in the *Cul3*^+/-^ mice.^15^

In this study, we describe a cohort of patients (n = 35), the largest to our knowledge, with *CUL3*-associated syndromic ID with autistic features. Through meta-analysis in large ASD and developmental delay (DD) trio-exome cohorts (n = 46,247), functional studies using patient derived cells, and facial profiling, we provide additional evidence supporting the role of *CUL3* in ASD and/or ID with variable penetrance. Our study helps to identify and further define the clinical spectrum, including shared syndromic features among patients with LoF *CUL3* variants.

## Materials and Methods

### Identification of pathogenic *CUL3* variants

We collected pathogenic variants in *CUL3* from exome, genome, or gene panel sequencing studies in patients through diagnostic clinical practice or Institutional Review Board approved research studies. Research subjects were also identified through GeneMatcher,^19^ Deciphering Developmental Disorders (DDD) cohort,^20^ and other professional communications. All variant coordinates are reported on GRCh37 (hg19) and transcript NM_003590.4.

### Patient consent

Patient consent for participation and phenotyping was obtained through each of the referring clinicians and centers. Clinical records including information related to neurodevelopmental, behavioral, dysmorphology, and other related phenotypic features were collected. Consent for collection of patient blood samples and clinical information, including patient photos, was further obtained in accordance with the recognized standards of the Declaration of Helsinki and approved by local Institutional Review Boards at the referring institutions.

### Statistical analyses

We collected six large exome and genome sequencing cohorts, excluding potential sample duplicates. For example, for cohorts including the DDD study, Autism Sequencing Consortium (ASC), and Radboud cohort, we only included the samples in their latest publication,^16^ where potential duplicates have been removed as described in their study methodology. We also excluded all the Simons Simplex Collection (SSC) samples used in the ASC study^17^ to avoid any potential redundancy with the SSC exomes.^11^ We performed *de novo* enrichment analyses using three models: the chimpanzee-human (CH) divergence model, denovolyzeR, and DeNovoWEST. Each method applies its own underlying variant rate estimates to generate the prior probabilities for observing a specific number and class of variants for a given gene: 1. CH model estimates the number of expected *de novo* variants by incorporating locus-specific transition, transversion, and indel rates, as well as chimpanzee–human coding sequence divergence and gene length; 2. denovolyzeR estimates mutation rates based on trinucleotide context and incorporates exome depth, mutational biases such as CpG hotspots, and divergence adjustments based on macaque–human comparisons; 3. DeNovoWEST scores all classes of coding variants on a unified severity scale based on the empirically-estimated positive predictive value of being pathogenic. It incorporates a gene-based weighting derived from the deficit of protein-truncating variants in the general population and further combines missense enrichment by a clustering test. Default parameters were used for all three methods as described previously.^16,18^ The expected mutation rate of 1.8 *de novo* variants per exome was set to the CH model as an upper bound baseline. The exome-wide significance cutoff (p-value < 4.16E-07) is determined after the Bonferroni multiple-testing correction considering both the ∼20,000 genes in a human genome and the total of six tests per gene across the three models.

### Patient facial analysis

We utilized GestaltMatcher^21^ to measure the similarities of the facial phenotypes between individuals with variants in *CUL3*. The feature vectors derived from GestaltMatcher were utilized to span a 512-dimensional clinical face phenotype space (CFPS). Each image is a point located in CFPS, and the similarity between the two images was quantified by the cosine distance. In CFPS, the images with close distance were considered to have a high overlap of syndromic facial features. To further improve the performance and stability, we utilized test-time augmentation and model ensemble techniques.

We first performed the statistical analysis to quantify the overall similarity among the 13 available facial photos (P1, P2, P3, P19, P20, P22, P23, P24, P25, P28, P29, P30, and P31) and compared to the control distributions. 1,696 images of 1,335 patients and 328 syndromes of the GestaltMatcher Database (GMDB; https://db.gestaltmatcher.org) were used to derive control distributions of mean pairwise cosine distances in CFPS stemming from a) patients with the same syndrome and b) random patients with different syndromes. For each syndrome, 100 cohorts of random sample sizes were drawn, and the mean pairwise cosine distance of the patients was calculated. Only unique cohorts were used to construct the distribution of mean pairwise cosine distance of patients stemming from the same syndrome. Additionally, the same number of cohorts consisting of random patients was drawn and used to construct the distribution of mean pairwise cosine distances of random patients stemming from different syndromes. Via a ROC analysis, a threshold to distinguish cohorts stemming from the same syndrome and random patients is derived by maximizing the Youden-index (sensitivity+specificity-1). We applied this approach in a five-fold cross-validation, and the validation fold was used to test the Youden-index of the resulting threshold. Finally, the control distributions and the corresponding threshold with the highest Youden index on the validation fold was chosen.

To test whether patients from a given cohort share a common phenotype, their mean pairwise cosine distance is calculated and compared to the derived threshold. Because P19 and P20 are family members, and P24 and P25 are also family members, the distances between related individuals are discarded to avoid the confounder. Additionally, 100 random subcohorts were drawn to estimate the uncertainty of the mean pairwise cosine distance. If a proportion of at least 50% of those mean pairwise cosine distances falls below the threshold, it is considered evidence for a shared phenotype within the cohort.

To further investigate the similarity among patients on the individual level, we performed the pairwise comparison analysis on 13 *CUL3* patients and compared them to 4,306 images with 257 different disorders from GMDB to simulate the real-world scenario. For each test image of a CUL3 patient, we performed the leave-one-out cross-validation by putting the rest 12 *CUL3* patients into the space with 4,306 images and calculating the ranks of 12 patients to the testing image.

### Patient T cells

Peripheral blood mononuclear cells (PBMCs) used in this study were isolated from whole blood from patients and the related healthy family members (father and/or mother of the proband) by PBMC spin medium gradient centrifugation (pluriSelect). The collected PBMCs were expanded in U-bottom 96-well plates using feeder cells in RPMI 1640 supplemented with 10% human AB serum (both purchased from PAN-Biotech GmbH) with 150 U/ml IL-2 (Miltenyi Biotec) and 1 µg/µl L-PHA (Sigma) following the procedure of Fonteneau et al.^23^ After 3-4 weeks of culture, resting T cells were washed and frozen as dry pellets for further use.

### SDS-PAGE and western blot analysis

Cell pellets from resting T cells isolated from patients and related controls were subjected to protein extraction by resuspending them in equal amounts of standard RIPA buffer (50 mM Tris pH 7.5, 150 mM NaCl, 2 mM EDTA, 1 mM N-ethylmaleimide, 10 µM MG-132, 1% NP40, 0.1% SDS) before separation by 10 or 12.5% SDS-PAGE and subsequent transfer to PVDF membranes (200V for 1h). Primary antibodies used in this study include anti-pan ubiquitin (Enzo Life Sciences, clone FK2), anti-ubiquitin K48-linkages (Cell Signaling Technology, clone D9D5), anti-CUL3 (Elabscience®, E-AB-10263), anti-cyclin E1 (Cell Signaling Technology, clone D7T3U), anti-CDK2 (Cell Signaling Technology, clone 78B2), anti-cyclin A2 (Cell Signaling Technology, BF683), anti-4E-BP1 (Cell Signaling Technology, clone 53H11), anti-GAPDH (Cell Signaling Technology, clone 14C10) and anti-α-tubulin (Abcam, clone DM1A). Proteins were visualized using anti-mouse or –rabbit HRP conjugated secondary antibodies (1/5,000) and an enhanced chemiluminescence detection kit (ECL) (Bio-Rad).

### RNA isolation, reverse-transcription and PCR analysis

Blood specimens were collected by using the PaxGene Blood RNA System (Qiagen) to stabilize total RNA which was subsequently isolated using the PaxGene Blood RNA kit (Qiagen) following the manufacturer’s instructions. Total RNA (100-500 ng) was subjected to reverse-transcription using the M-MLV reverse transcriptase from Promega and qPCR was performed using the Premix Ex Taq™ (probe qPCR purchased from TaKaRa) in duplicates to determine the mRNA levels of each target gene using FAM-tagged TaqMan™ Gene Expression Assays (Thermo Fisher Scientific). TaqMan™ probes used in this study for Interferon-Stimulated Gene (ISG) quantification included *IFI27*, *IFI44L*, *IFIT1*, *ISG15*, *RSAD2*, and *SIGLEC1*. The cycle threshold (Ct) values for target genes were converted to values of relative expression using the relative quantification (RQ) method (2-ΔΔCt). Target gene expression was calculated relative to Ct values for the *HPRT1* control housekeeping gene. A type I IFN score was calculated for each sample following the procedure of Rice et al.^24^

## Results

### Excess of *de novo* variants in *CUL3* among a large NDD cohort

Among our recent collection of *de novo* variants from a large NDD cohort with 46,247 trios (15,189 ASD and 31,058 DD)^11,16,17, 26–28^, there are three *de novo* missense variants and 16 *de novo* LoF variants (including 10 frameshift, 3 stop-gain, 2 splice-donor and 1 splice-acceptor) identified in *CUL3* (Table S1, Table S2). We performed a comprehensive *de novo* enrichment analysis across all *de novo* variants and genes among this collection, where three different statistical models were applied independently in parallel (CH model,^9,25^ denovolyzeR,^29^ and DeNovoWEST^16^). After Bonferroni correction, exome-wide significance was supported by all three models for excess of *de novo* variants in *CUL3* (p = 4.60E-24 in CH model, p = 2.59E-17 in denovolyzeR, p = 6.8E-11 in DeNovoWEST). Our observations are in line with the intolerant scores in gnomAD v2.1.1,^30^ which show that *CUL3* is highly intolerant to both missense and LoF (truncating) variants, with a missense Z score of 4.75 (observed/expected ratio [o/e] = 0.35 [0.3 - 0.4]) and the probability of being LoF intolerant (pLI) being 1 (o/e = 0.1 [0.05 - 0.23]). Those two lines of evidence together further strengthen the role of *CUL3* variants in NDD.

### Molecular findings in the newly described *CUL3* clinical series

Via a multi-center collaboration, we collected 35 individuals with rare heterozygous variants in *CUL3* (Table 1), including 33 LoF variants (17 frameshift, 10 nonsense, and 6 with canonical splice site variants) and two missense variants. Of these, 22 are *de novo*, nine are inherited, and inheritance was unknown for the remaining four individuals as DNA from both parents was unavailable. Parental inheritance was noted in families F7, F8, F11, F17, F18, F22, and F25 (Fig. 1). Formal evaluations and clinical details are provided for P20 (mother of P19 in F18) and P25 (mother of P24 and P26 in F22). Limited clinical information was available for several reportedly affected parents (Mother of P7 and P8 in F7; Father of P12 in F11). The mother of P9 (F8) has reported learning difficulties but was not considered to have ID. The mother of P18 (F17) was reported to have mild ID with some speech difficulties, but a full evaluation was not performed. Although the mother of P29 (F2) is reported to be unaffected, she has not received a formal evaluation in the clinical setting. Recurrent LoF variants were observed in multiple unrelated families. Specifically, p.Ser517Profs*23 had the highest incidence and was observed in four unrelated individuals (2 *de novo* and 2 inherited), and p.Leu165Ilefs*37 was identified in two unrelated individuals (both *de novo*).

**Figure 1.**
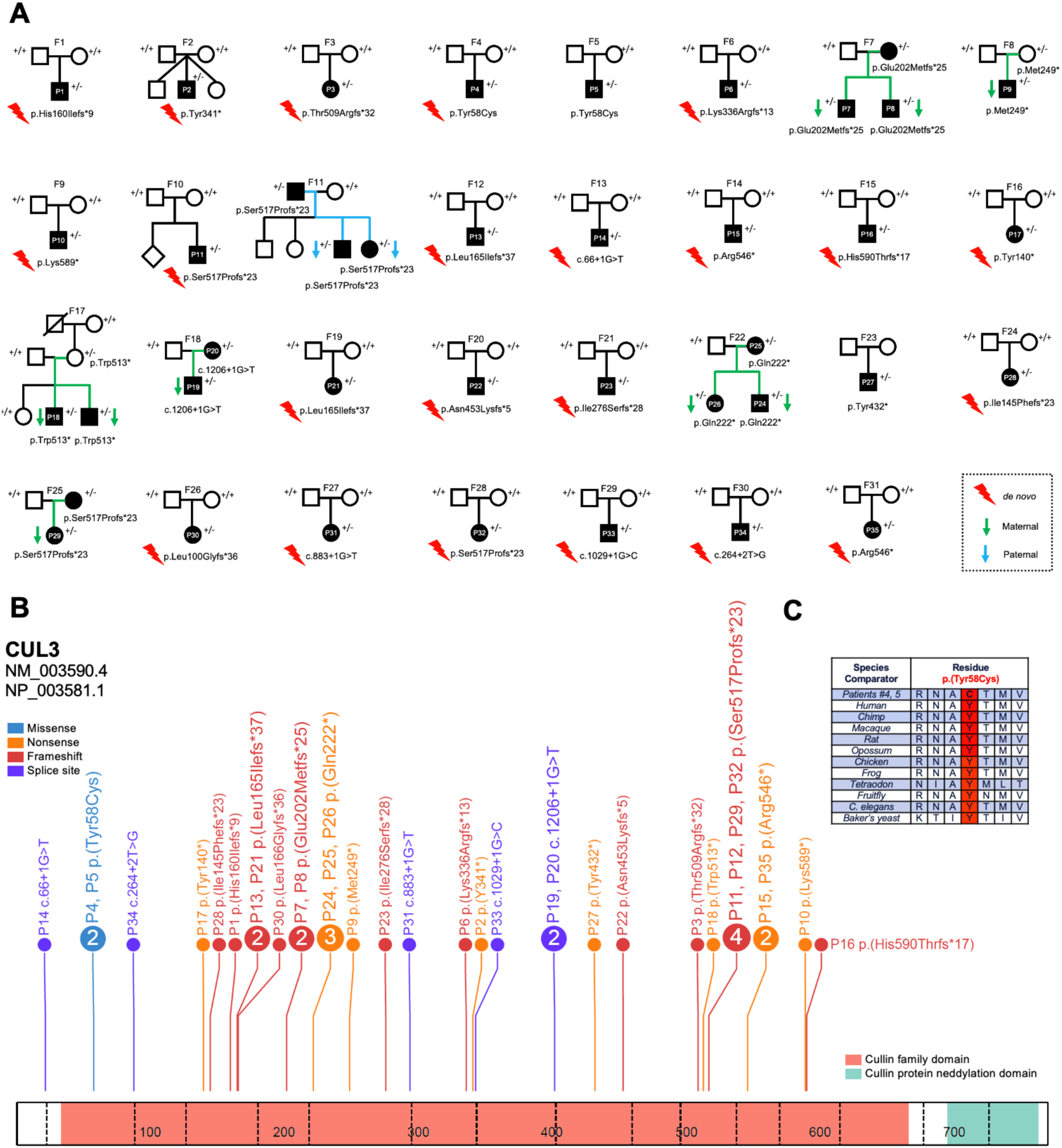
Samples in study. A) Families and patients with index number included in study. Variants with a symbol of red lightning bolt are *de novo*, maternally inherited variants are indicated with a green arrow and paternally inherited variants with blue arrow. B) Variant distribution on the protein diagram of CUL3. C) Amino acid conservation across species for the missense variant (p.Tyr58Cys) in P4 and P5.

**Table 1.** Patients and variants in study.

| Family ID | Patient ID | Sex * | HGVSc | HGVSp | Variant | Inheritance | CADD score (v1.3) |
| --- | --- | --- | --- | --- | --- | --- | --- |
| F1 | P1 | M | c.478del | p.His160Ilefs*9 | frameshift | *de novo* | - |
| F2 | P2 | M | c.1022_1023insG | p.Tyr341* | stop_gained | *de novo* | - |
| F3 | P3 | F | c.1526del | p.Thr509Argfs*32 | frameshift | *de novo* | - |
| F4 | P4 | M | c.173A>G | p.Tyr58Cys | missense | *de novo* | 27.9 |
| F5 | P5 | M | c.173A>G | p.Tyr58Cys | missense | Not maternal | 27.9 |
| F6 | P6 | M | c.1007del | p.Lys336Argfs*13 | frameshift | *de novo* | - |
| F7 | P7 | M | c.602_603dup | p.Glu202Metfs*25 | frameshift | Maternal (mother affected, brother of P8) | - |
|  | P8 | M | c.602_603dup | p.Glu202Metfs*25 | frameshift | Maternal (mother affected, brother of P7) | - |
| F8 | P9 | M | c.743_744dup | p.Met249* | stop_gained | Maternal | - |
| F9 | P10 | M | c.1765A>T | p.Lys589* | stop_gained | *de novo* | 41 |
| F10 | P11 | M | c.1549_1552del | p.Ser517Profs*23 | frameshift | *de novo* | - |
| F11 | P12 | F | c.1549_1552del | p.Ser517Profs*23 | frameshift | Paternal (father affected) | - |
| F12 | P13 | M | c.493_494del | p.Leu165Ilefs*37 | frameshift | *de novo* | - |
| F13 | P14 | M | c.66+1G>T | - | splice_donor | *de novo* | 34 |
| F14 | P15 | M | c.1636C>T | p.Arg546* | stop_gained | *de novo* | 41 |
| F15 | P16 | M | c.1767_1768insA | p.His590Thrfs*17 | frameshift | *de novo* | - |
| F16 | P17 | F | c.420C>A | p.Tyr140* | stop_gained | *de novo* | 37 |
| F17 | P18 | M | c.1538G>A | p.Trp513* | stop_gained | Maternal (mother unaffected) | 43 |
| F18 | P19 | M | c.1206+1G>T | - | splice_donor | Maternal (mother P20 affected) | 36 |
|  | P20 | F | c.1206+1G>T | - | splice_donor | Unknown (affected mother of P19) | 36 |
| F19 | P21 | F | c.493_494del | p.Leu165Ilefs*37 | frameshift | *de novo* | - |
| F20 | P22 | M | c.1358dup | p.Asn453Lysfs*5 | frameshift | *de novo* | - |
| F21 | P23 | M | c.827_828del | p.Ile276Serfs*28 | frameshift | *de novo* | - |
| F22 | P24 | M | c.664C>T | p.Gln222* | stop_gained | Maternal (mother P25 affected) | 39 |
|  | P25 | F | c.664C>T | p.Gln222* | stop_gained | Unknown (affected mother of P24 & P26) | 39 |
|  | P26 | F | c.664C>T | p.Gln222* | stop_gained | Maternal (mother P25 affected) | 39 |
| F23 | P27 | M | c.1296T>G | p.Tyr432* | stop_gained | Unable to confirm status (singleton exome) | 37 |
| F24 | P28 | F | c.433_436del | p.Ile145Phefs*23 | frameshift | *de novo* | - |
| F25 | P29 | F | c.1549_1552del | p.Ser517Profs*23 | frameshift | Maternal (mother unaffected; no formal evaluation) | - |
| F26 | P30 | F | c.297_298del | p.Leu100Glyfs*36 | frameshift | *de novo* | - |
| F27 | P31 | F | c.883+1G>T | - | splice_donor | *de novo* | 33 |
| F28 | P32 | F | c.1549_1552del | p.Ser517Profs*23 | frameshift | *de novo* | - |
| F29 | P33 | M | c.1029+1G>C | - | splice_donor | *de novo* | 35 |
| F30 | P34 | M | c.264+2T>G | - | splice_donor | *de novo* | 33 |
| F31 | P35 | F | c.1654C>T | p.Arg552* | stop_gained | *de novo* | 41 |

*CUL3* is highly conserved among orthologues from several species, and the residues affected by missense variation show a high degree of evolutionary conservation (p.Tyr58Cys in unrelated individuals, P4 and P5; conserved in baker’s yeast, *Saccharomyces cerevisiae*) (Fig. 1). These variants are predicted to be damaging by multiple tools (CADD, PolyPhen2, SIFT). Additionally, all variants are absent or extremely rare in the population (gnomAD v2.1.1) (Table S3, Table S4).^30^ All five canonical splice site variants were predicted to result in donor loss and/or altered splicing of several exons and were notably absent from exon 9, which is affected in patients with pseudohypoaldosteronism type IIE (Fig. 1, Table S5).

Three patients had secondary genetic findings of uncertain clinical significance, and their contribution to the overall clinical phenotype in these individuals is unclear. P2 carries a *de novo* nonsense variant (p.Y341*) in *CUL3* and a *de novo* missense variant in *CHD3* [NM_001005273.2, c.5863G>T, p.A1955S]. The variant in *CHD3* was previously reported in Drivas et al.^31^, falls near the end of the protein in an unannotated portion of the protein, and does not cluster with other disease-implicated missense variants in the conserved SNF2 family N-terminal or Helicase C-terminal domains. P31 has a *de novo* splice site variant (c.883+1G>T) in intron 6 of *CUL3*, together with a *de novo* missense variant (NM_001282948.1, c.6260A>C, p.Asp2087Ala) in *JMJD1C*. *De novo* heterozygous missense and LoF variants in *JMJD1C* have been implicated variably in patients with DD, ASD, and a phenotype resembling classical Rett syndrome.^33,34^ P32 has a *de novo* frameshift variant (p.Ser517Profs*23) in *CUL3*, as well as a 22q12.3 duplication [arr 22q12.3(33,421,825-34,080,737)x3 pat] that includes a portion of the *LARGE* and *SYN3* genes, which was inherited from his apparently normal father. Overlapping duplications have been implicated in increased risk of ASD and other NDDs, however the significance of this relatively small duplication is still unclear.^35^ To our knowledge, no other patients had secondary reportable alterations of clinical significance in this cohort.

### Variants in *CUL3* define a neurodevelopmental syndrome

The 35 cases (22 males and 13 females) included in the study are from 31 families who carry heterozygous, mainly predicted LoF variants in *CUL3* through genome, exome, or custom gene panel sequencing. Affected individuals presented with global DD (94%, 33/35), delayed speech and language development (97%, 32/33), dysmorphic facial features (high forehead, long face, and other variable features) (93%, 28/30) (Fig. 2A), mild to severe ID (90%, 28/31), gross and fine motor delays (67%, 18/27 and 96%, 27/28, respectively), ASD (38%, 13/34), abnormalities of the hands and feet (55%, 18/33) (Fig. 2B) (including bilateral 5^th^ finger clinodactyly, thenar hypoplasia, single palmar crease, ankle/foot contractures, pes cavus, cutaneous syndactyly second/third toes, hallux valgus etc.), and tremor and/or dystonia (20%, 7/35) (Table 2 and Table S4).

**Figure 2.**
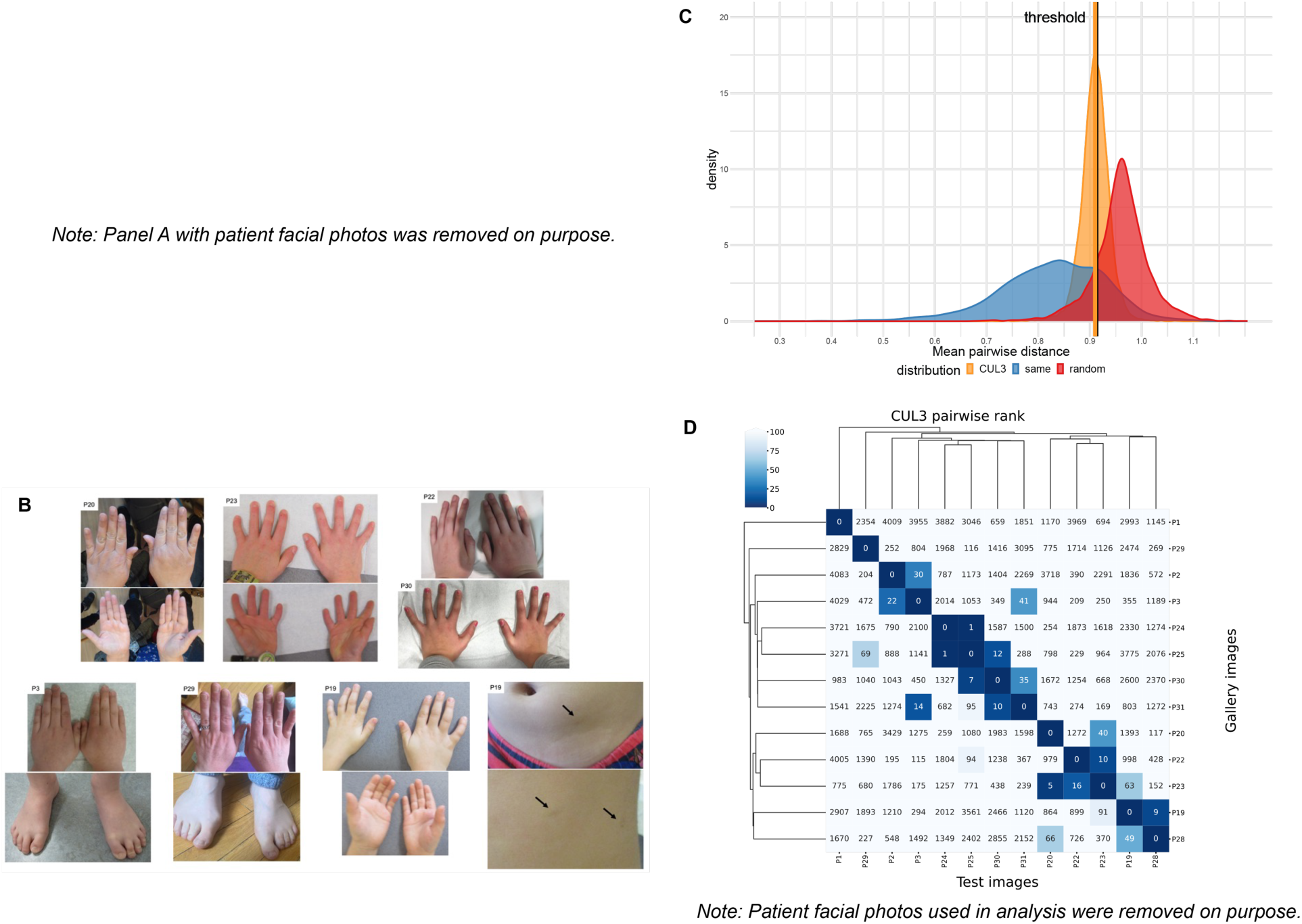
The patient phenotype, distribution of mean pairwise distance, and the hierarchical clustering of facial photos. The phenotypic photos for available patients are shown in A) and B). C) It shows three distributions: *CUL3* (orange), the random selection from the subjects with 328 disorders (red), and the selection with the same disorder (blue). 53.72% of the *CUL3* sampling with mean pairwise distance (0.9100) were below the threshold (0.9124; blackline). D) Gallery images were the images in clinical face phenotype space (CFPS) which can be matched. Each column is the result of testing one subject in the column and listing the rank of the rest for the 12 photos in each row. For example, by testing P25, P30 was on the 7th rank and P23 was on the 5th rank of P20.

**Table 2.**
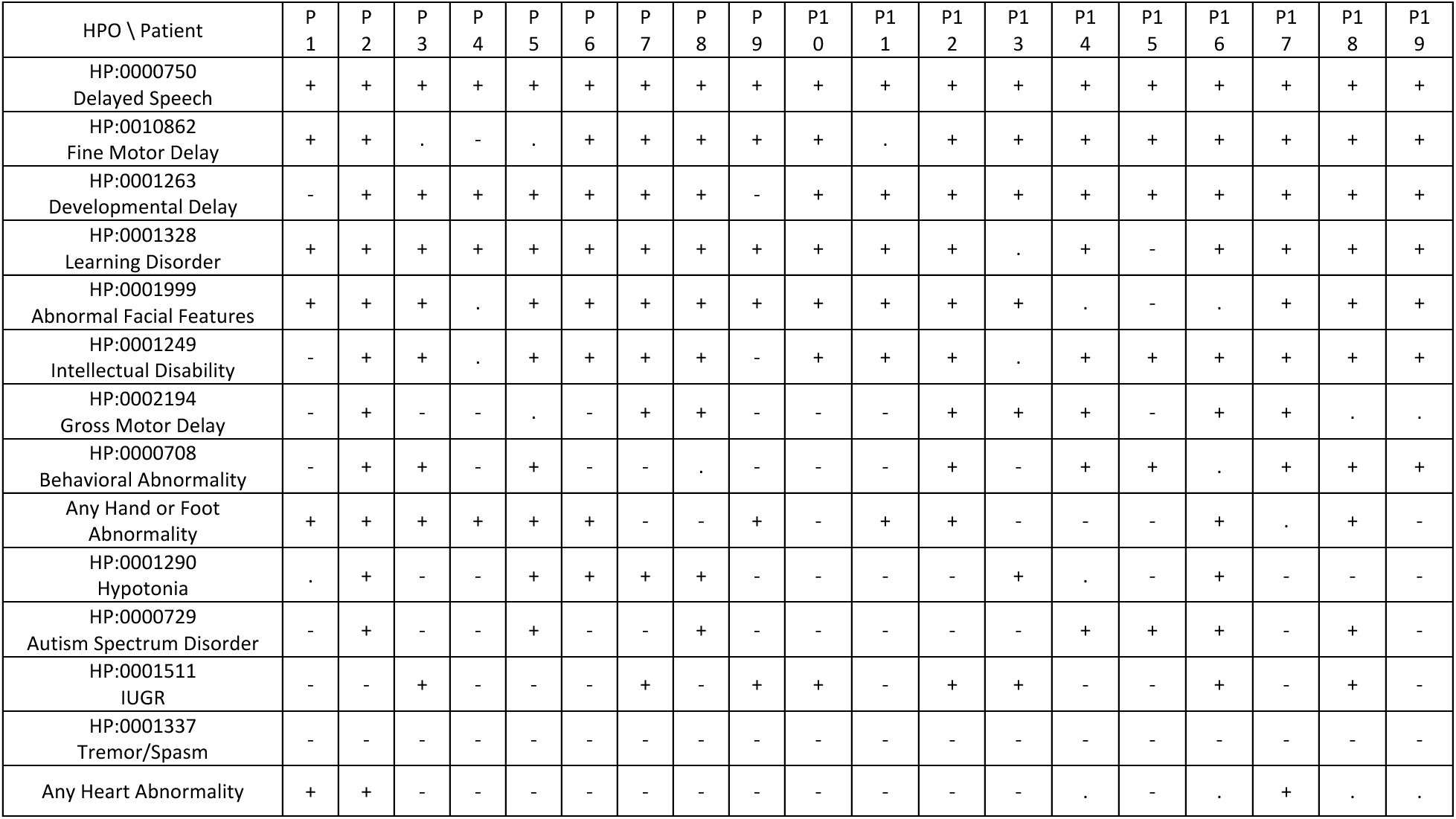
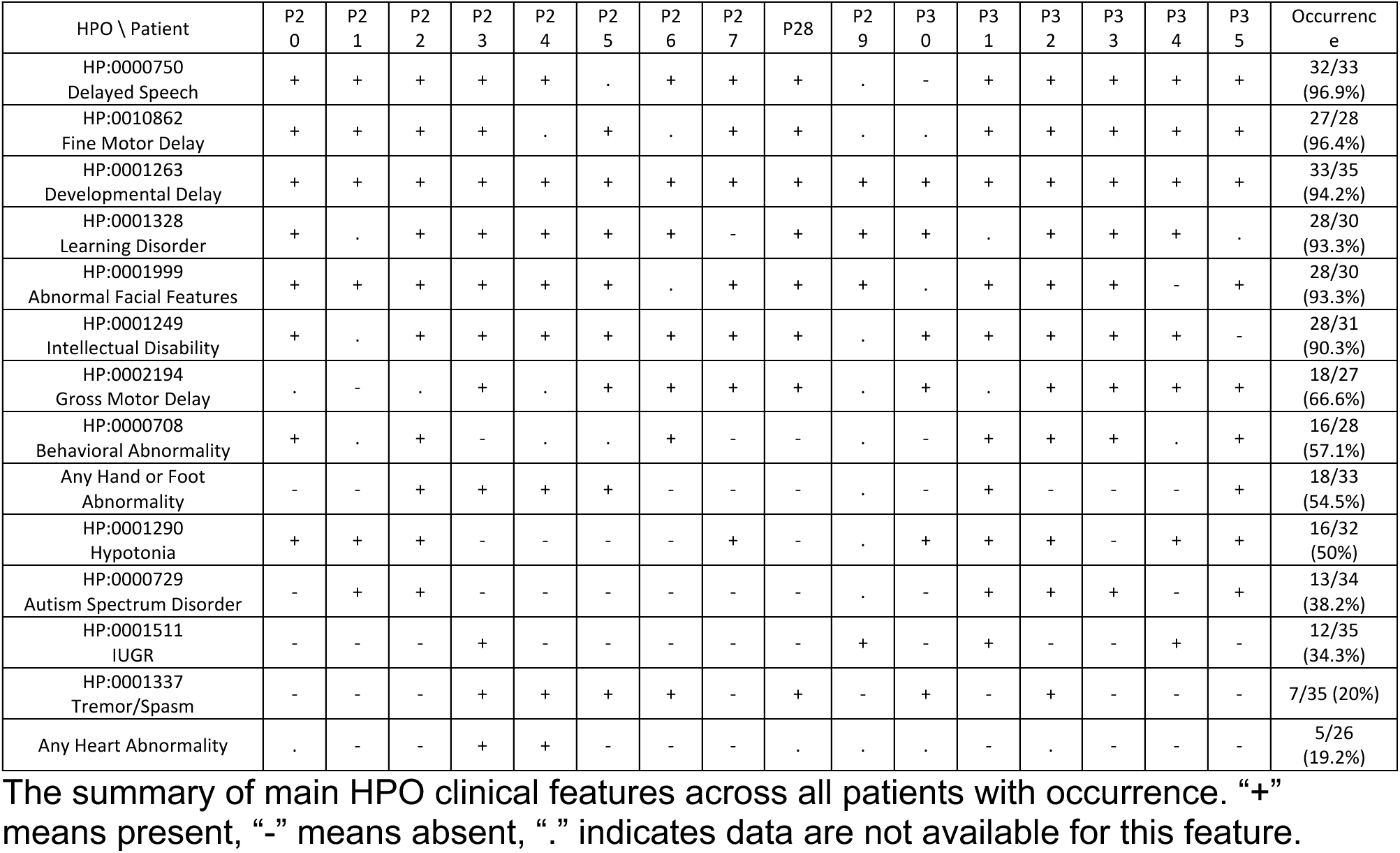
Summary of clinical features.

### Prenatal and early postnatal presentation

Approximately 34% (12/35) of patients were found to have intrauterine growth restriction (IUGR) during gestation. Birth length, weight, and occipitofrontal circumference (OFC), when available, were variable but usually fell within the low-normal range (Table S4). Gastroesophageal reflux disease (GERD) was noted in infancy in 33% (9/27) of subjects. Cleft palate was observed in one individual, and cryptorchidism was the most common genitourinary abnormality noted at birth (3 individuals).

### Facial features

As noted above, most individuals in the cohort were reported to have dysmorphic facial features, which were usually mild and were somewhat variable (Fig. 2A). Several individuals were macrocephalic, with long triangular facies, including a large forehead and pointed chin. Ears were slightly dysplastic to normal and were small and posteriorly rotated in some individuals. The nose was bulbous in some with a wide nasal bridge. Some patients had a high arched palate and/or retrognathia. Eyes were deep set with slightly narrow/downslanting palpebral fissures with epicanthal folds noted in some. Strabismus was noted in four individuals.

To further study defining facial features, we performed analyses with GestaltMatcher using individual photographs of the 13 patients for whom photographs were available. GestaltMatcher can be used to validate whether patients in a given cohort share a common phenotype. To do so, the mean pairwise cosine distance between patients of a cohort is analyzed. It was then compared to two control distributions of mean pairwise cosine distances stemming from a) patients with the same syndrome and b) random patients with different syndromes and a threshold c to distinguish cohorts stemming from a) or b) can be derived, resulting in c=0.9124 with a corresponding sensitivity of 0.86 and a specificity of 0.79. For CUL3, we calculated the mean pairwise distance of 13 patients while distances between related individuals were discarded. This resulted in a mean pairwise cosine distance d=0.910<c. Furthermore, 53.72% of mean pairwise cosine distances of sampled sub-cohorts fell below the threshold (Fig. 2C). Overall, the analysis supports the hypothesis of a shared phenotype among *CUL3* patients.

To further investigate the similarity on the individual level, pairwise ranks analysis (Fig. 2D) was performed. Most of the individuals showed significant similarity as expected. Seven pairs of subjects, (P2, P3), (P3, P31), (P19, P28), (P20, P23), (P22, P23), (P25, P30), (P30, P31), were among the top-30 rank of each other in overall 4,306 subjects. Moreover, P30 was at the 7^th^ rank of P25, P23 was at the 5^th^ rank of P20. Overall, the results showed a high degree of facial similarity, suggesting a recognizable pattern of facial dysmorphism. Of note, previous studies have shown that DeepGestalt can be influenced by confounding factors such as age and ethnic background.^36,37^ In the clustering dendrogram, the pair of P24 and P25 were at the 1^st^ rank to each other. It could be due to confounding factors since they are family members. A more comprehensive analysis of the confounding effects is required when more photos are collected in the future.

### Developmental delay and intellectual disability

Speech delay was the most consistent feature and was noted in 97% (32/33). It was usually mild but moderate to severe in seven individuals. Some form of ID was noted in 90% of patients (28/31), though several patients were too young to assess. The severity of ID was again mild in most cases and moderate to severe in nine cases. Formal IQ testing was performed in eight individuals and ranged from 49 to 108 (median 62). Early hypotonia was seen in 50% of individuals (16/32) but improved or resolved with age in most individuals. There was a high degree of clinical suspicion for or a formal diagnosis of ASD in 38% of patients (13/34). Additional abnormalities and behavioral issues were noted in 57% of patients (16/28) and consisted of dysgraphia, slow information processing and spatial reasoning, agitation, (hetero) aggressivity, impaired social interactions, poor frustration tolerance, and OCD and repetitive behaviors. No consistent brain structural abnormalities were noted in the patient cohort. Most individuals who had a brain MRI performed had a normal study, but three were noted to have mild ventriculomegaly. Seizures, in contrast to previous studies, were observed in only a few patients and were either febrile seizures or consisted of single isolated episodes suggesting that this may not be a recurrent clinical feature.

### Dystonia and movement abnormalities

Movement abnormalities were noted in several patients (7/35 with noted tremor/spasm) in this cohort including five individuals with variable dystonia. P23 had *de novo* p.Ile276Serfs*28 and developed tremor, myoclonus, and had severe motor delays. P24 and P26 both carried a maternally inherited variant (p.Gln222*); P24 had hyperreflexia, hand ataxia, cervical dystonia (onset at ∼20-25 years of age), truncal dystonia and laryngeal dystonia (onset at ∼20-25 years of age) which led to speech problems. P26, the affected female sibling of P24, was diagnosed with diparetic form of cerebral palsy, with a history of congenital hydrocephalus, and temporal epilepsy. Their mother, P25, had hand tremor since childhood, which was not very progressive, adult onset oromandibular dystonia with speech involvement, and sudden recent onset of hyperreflexia. P28, with *de novo* p.Ile145Phefs*23, had generalized dystonic body movements and spasms in her lower limbs (onset at ∼25-30 years of age); her episodes of muscle spasms were associated with shortness of breath, sweating, high blood pressure and tachycardia. Finally, P30 (*de novo* p.Leu100GlyfsTer36) developed facial hypotonia during infancy and later left inferior limb dystonia, with slow movements and an unusual gait marked by bilateral hip internal rotation. This appears to be a novel finding in this cohort of patients, and further study is warranted.

### Hypertension and other cardiac abnormalities

Several LoF mutations in *CUL3* have been reported in large trio exome studies of congenital heart disease.^7,38^ One patient with a *de novo* frameshift variant (p.Ile145Phefs*23) in *CUL3* was described with left ventricular obstruction, hypoplastic mitral valve, hypoplastic aortic annulus, aortic stenosis, coarctation, congenital hip dysplasia, and congenital scoliosis.^7^ Another patient with *de novo* stop-gain variant (p.Gln132*) had a left aortic arch with normal branching pattern, partially anomalous pulmonary veins, pulmonary valve stenosis, and sinus venosus septal defect, superior type.^38^ In this study, we found three patients with various heart abnormalities: P1 with pulmonic stenosis and pulmonary valve dysplasia, P17 with a ventricular septal defect, and P24 with mild mitral valve insufficiency (ejection fraction 55%, cardiac MRI normal). Two patients presented with conduction defects including one with sinus rhythm with short PR interval and nonspecific T wave abnormalities, and one with sinus rhythm and supraventricular and rare ventricular extrasystoles. Five patients presented with unexplained early onset/juvenile hypertension in the absence of hyperkalemia.

### *CUL3* variants are associated with ubiquitin conjugation deficiency and accumulation of CUL3 substrates in patient cells

Because CUL3 serves as a scaffolding subunit of an E3 ubiquitin ligase complex supporting the ubiquitination of several substrates,^39^ we next evaluated the functional consequences of the identified *CUL3* alterations on the ubiquitin-conjugation system in patient cells. In total, T cells were isolated from six individuals carrying four *CUL3* variants: p.Tyr58Cys (P4 and P5), p.Ser517Profs*23 (variant observed in 4 unrelated individuals [P11, P12, P29, and P32]), p.Trp513* (proband P18 and his unaffected mother), or c.1206+1G>T (proband P19 and his mother P20) were assessed for their content in ubiquitin-modified proteins by western-blotting using an anti-pan ubiquitin antibody (FK2). The amounts of ubiquitin-protein conjugates were substantially reduced in all investigated patients when compared to those from a healthy donor control (Fig. 3A). Probing the membrane with an antibody specific for ubiquitin K48 linkages revealed a similar pattern, indicating that patients failed to assemble ubiquitin chains for the modification of substrates destined for proteasomal degradation. Importantly, the decreased ubiquitination detected in patients with *CUL3* alterations was accompanied by a decreased steady-state expression of the CUL3 full-length protein (Fig. 3B). Of note, we could not detect the truncated variants, p.Ser517Profs*23 and p.Trp513*, with a predicted size of 63 and 60 kDa, respectively. Likewise, our anti-CUL3 antibody failed to detect any truncated CUL3 or aberrantly spliced species emerging from the c.1206+1G>T alteration (Fig. 3B).

**Figure 3.**
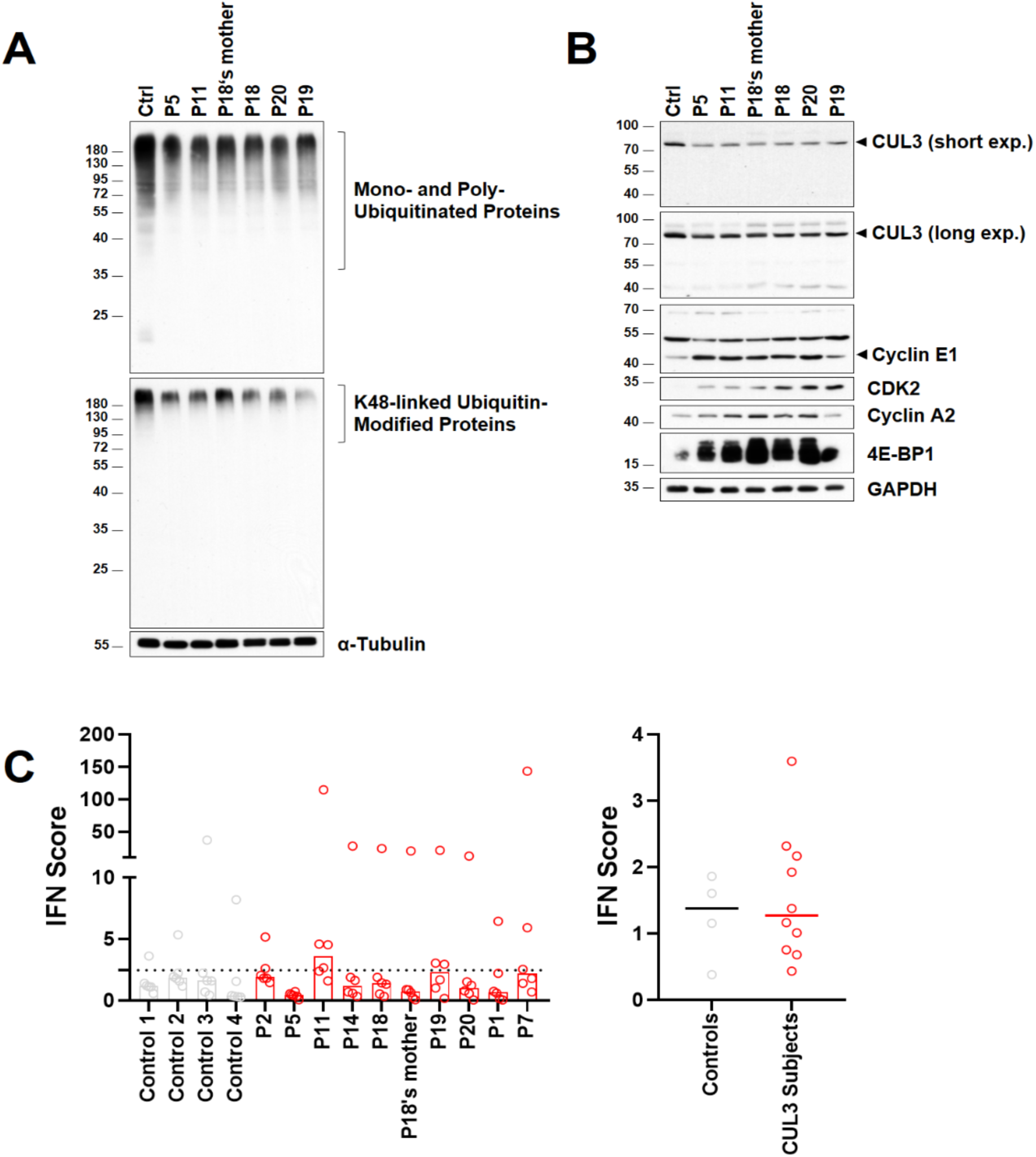
Cells isolated from subjects carrying *CUL3* variants exhibit protein homeostasis perturbations resulting in the stabilization of CUL3 substrates. A) Five micrograms of RIPA lysates from T cells isolated from patients with heterozygous pathogenic *CUL3* variants and controls (Ctrl) were separated by SDS-PAGE followed by western blotting using antibodies directed against pan-ubiquitin and K48-linked ubiquitin-modified proteins, as indicated. Equal protein loading was ensured by probing the membrane with an anti-α-tubulin antibody. B) Five to forty micrograms of whole-cell lysates were assessed for their CUL3, cyclin E1, CDK2, cyclin A2, 4E-BP1, and GAPDH (loading control) protein content, as indicated. C) *CUL3* loss of function is not associated with a type I IFN gene signature. IFN scores for the CUL3 subjects (controls and patients) were calculated as the median of the relative quantification (RQ) of the six interferon-stimulated genes (ISGs) over a single calibrator control. Shown are the IFN scores of each sample (left panel) and the sample groups, i.e., controls and *CUL3* subjects, as indicated (right panel).

Since CUL3 has been shown to target cyclin E1 and 4E-BP1 for degradation,^40,41^ we next examined the expression level of both proteins in patient cells. A significant stabilization of these substrates was detected in patient-derived T cells, thereby confirming the inability of CUL3 to ubiquitinate and target them for proteasomal degradation (Fig. 3B). Taken together, these data point to a clear role of *CUL3* LoF variants in patient cells impacting the protein turnover of several components of the cell cycle and translational machinery.

### *CUL3* LoF variants do not promote a type I IFN signature

Because proteasome LoF variants typically associated with the generation of a sterile type I IFN response in patients with proteasome-associated autoinflammatory syndromes (PRAAS),^42–48^ we next attempted to clarify whether *CUL3* variants generate such a signature as well. To address this, we compared the expression of six IFN-stimulated genes (i.e. *IFI27*, *IFI44L*, *IFIT1*, *ISG15*, *RSAD2* and *SIGLEC1*) in PAXgene blood RNA samples from patients and controls. Our data revealed that the IFN scores did not substantially differ between patients and controls, indicating that *CUL3* LoF variants, despite disrupting protein homeostasis of several known targets, do not promote a type I IFN gene signature (Fig. 3C).

## Discussion

In this study, we defined the phenotypic spectrum in 35 individuals carrying rare heterozygous variants in *CUL3*. Except two patients with a recurrent missense variant observed in a highly conserved residue, the vast majority are frameshift, nonsense, or canonical splice site variants that are predicted to result in *CUL3* haploinsufficiency. The clinical manifestation, while variable, has a number of consistent findings including DD and ID in the vast majority of patients. Behavioral difficulties were also very prevalent with a subset diagnosed with ASD. Learning difficulties were most often accompanied with speech delay and other processing issues and ranged in severity from mild to profound. Patients also presented with both gross and fine motor delays early in development. A subset of patients presented with movement abnormalities including early hypotonia in addition to later (usually adult) onset dystonia and tremor in some individuals. However, epilepsy, which were thought to be a part of the clinical spectrum, were not observed in our patient cohort (OMIM #619239).

Several recurrent facial features were noted in our cohort including relative macrocephaly and a long triangular face with a prominent forehead and pointed chin (Fig. 2A). Ears were normal appearing in most individuals, but were small, dysplastic, and posteriorly rotated in some. Several individuals had a bulbous nose with a wide nasal bridge. Eyes were described as deep set with slightly narrow/downslanting palpebral fissures with epicanthal folds noted in several individuals. We performed facial analysis on 13 individuals with available frontal photos using GestaltMatcher, which identified several similarities among unrelated individuals in our cohort (Fig. 2C). In summary, most of the available subjects with *CUL3* variants presented with similar facial dysmorphisms. However, a comprehensive analysis with more photos to exclude possible confounding effects is required in the future.

*CUL3* is expressed in a variety of different tissues including brain and nerve tissue but has the highest expression levels in testis, skeletal muscle, heart, esophagus mucosa, skin, and the uterus (based on GTEx data; accessed on October 1^st^, 2022). We identified several clinical phenotypes affecting these tissues, suggesting that *CUL3* haploinsufficiency may influence expression of some of these traits. For instance, while the role of gain-of-function splice variants affecting exon 9 of *CUL3* is well known in renal hyperkalemia and hypertension in autosomal dominant pseudohypoaldosteronism, type IIE (OMIM# 614496), we identified several individuals in our cohort without electrolyte abnormalities who had early or juvenile onset hypertension or other congenital cardiac abnormalities. A few patients were noted to have abnormal (thickened) scarring, delayed wound healing, or easy bruising. One patient (P13) was noted to have cutaneous dyschromia (Fig. 2B). Although relatively nonspecific, several patients had gastroesophageal reflux disease in infancy and early childhood. Several additional congenital anomalies were noted, including abnormalities of the hands and feet (51.5%, 17/33) (Fig. 2B), most notably bilateral 5^th^ finger clinodactyly in 4 individuals, thin thumbs and thenar hypoplasia in two individuals, single palmar crease in 3 individuals, contractures involving the ankles, feet or toes in five (talipes equinovarus in 1 individual), pes cavus in five individuals, cutaneous syndactyly second/third toes in two individuals, and hallux valgus in two individuals. Several patients were noted to have scoliosis or severe lordosis. Oscillating testis, retractile testis, and cryptorchidism noted in one patient each. Overall the appearance of several recurrent clinical phenotypes in our cohort implicates *CUL3* in a syndromic ID/ASD disorder.

Functional studies showed that subjects harboring *CUL3* variants show severe signs of protein homeostasis perturbations, as evidenced by their decreased content of ubiquitin-protein conjugates (Fig. 3A). Such reduced amounts of ubiquitin-modified proteins might reflect a general ubiquitin-conjugation deficiency in these patients, as a consequence of loss of function for CUL3. In line with this assumption, cyclin E1 and 4E-BP1, two prominent substrates of CUL3, failed to be targeted for proteasomal degradation in all investigated subjects (Fig. 3B). The inefficiency of CUL3 to ubiquitinate its intracellular target proteins in patients might be due to insufficient expression levels, as evidenced by lower protein levels in mutant compared to control cells (Fig. 3B). Importantly, we were unable to detect any of the CUL3 truncated variants potentially resulting from the p.Ser517Profs*23, p.Trp513* and c.1206+1G>T alterations. One potential explanation for this result may be the specificity of our anti-CUL3 antibody, which may be exclusively directed to the C-terminal part of the protein and, as such, unable to recognize C-terminal truncated variants. Alternatively, it is also conceivable that such variants may be degraded by nonsense-mediated mRNA decay and therefore not subjected to translation. In any case, this issue warrants further investigation. The abnormal accumulation of cyclin E1 and its binding partner CDK2 in patients with *CUL3* variants (Fig. 3B) may actively contribute to the pathogenesis of the observed neurodevelopmental delay. Indeed, overexpression of cyclin E1 in embryonic murine brains has been associated with dysregulation of corticogenesis.^49^ Likewise, stabilized 4E-BP1 may be implicated in the neuronal phenotype by impeding protein translation.

Our data show that, unlike proteasome loss of function, *CUL3* LoF is not associated with a type I IFN gene signature (Fig. 3C). These data are interesting and suggest that the generation of such signature clearly depends on the type of protein homeostasis imbalance with which the cells are confronted. Hence, any shift towards accumulation of ubiquitin-modified proteins promotes a type I IFN response, while any change towards deficiency would not necessarily result in autoinflammation. Our report is currently the most comprehensive phenotypic analysis of patients with rare heterozygous mainly LoF variants in *CUL3*. Our cohort analysis suggests that *CUL3* haploinsufficiency is associated with a disorder characterized by ID with or without autistic features, variable congenital anomalies, and facial dysmorphisms, which together constitute a clinically recognizable syndrome. Our study also provides strong evidence supporting the role of CUL3 as a member of an emerging group of cullinopathies, which have wide-ranging effects on development, the biology of which is still being elucidated.

## Web Resources

Genome Aggregation Database (gnomAD), https://gnomad.broadinstitute.org/

ClinVar, https://www.ncbi.nlm.nih.gov/clinvar/

Combined Annotation Dependent Depletion (CADD), https://cadd.gs.washington.edu/

DECIPHER, http://decipher.sanger.ac.uk

GeneMatcher, https://genematcher.org/

GestaltMatcher, http://www.gestaltmatcher.org/

GTExPortal: https://gtexportal.org/

Mutation Taster, http://www.mutationtaster.org/

Online Mendelian Inheritance in Man (OMIM), http://www.omim.org/

PolyPhen2, http://genetics.bwh.harvard.edu/pph2/

ProteinPaint, https://proteinpaint.stjude.org/

Sorting Intolerant from Tolerant (SIFT), https://sift.bii.a-star.edu.sg/

SpliceAI, https://spliceailookup.broadinstitute.org/

Variant Score Ranker (VSR), http://vsranker.broadinstitute.org/

## Data Availability

All of the patient variant and phenotype data used in this study has been provided in the tables, or relevant supplementary tables.

## Acknowledgements

We would like to express our sincere gratitude to all the families for their participation in this research study. We thank Tonia Brown for assistance in editing this manuscript. This research was made possible through access to the data and findings facilitated by the web-based tools of GeneMatcher and DECIPHER. A full list of centers that contributed to data generation is available at http://decipher.sanger.ac.uk and via email at.

## Author Contributions

P.B., F.E., S.K., M.T., T.W. designed the study and supervised the work. F.E., T.C.H., and T.W. conducted analyses. All the other authors provided genetic and phenotypic data for the study. P.B., S.K., M.T. provided critical feedback on all analyses and figures. The paper was written by P.B., F.E., T.C.H. and T.W. with comments provided by all other authors.

## Ethics Declaration

The study was designed and initiated while P.B. was at the Mayo Clinic. Approval for the study was provided by the Mayo Clinic Institutional Review Board (IRB# 12-009346). F.E. performed functional investigations on human samples which were approved by the ethics board of the Universitätsmedizin Greifswald (Ethics number BB 014/14). Patient consent for participation and phenotyping was obtained through each of the referring clinicians and centers. Clinical records including information related to neurodevelopmental, behavioral, dysmorphology, and other related phenotypic features were collected. Consent for collection of patient blood samples and clinical information, including patient photos, was further obtained in accordance with the recognized standards of the Declaration of Helsinki and approved by local Institutional Review Boards at the referring institutions.

